# Assessing health equity in wastewater monitoring programs: Differences in the demographics and social vulnerability of sewered and unsewered populations across North Carolina

**DOI:** 10.1101/2023.10.06.23296680

**Authors:** Xindi C. Hu, Stacie K. Reckling, Aparna Keshaviah

## Abstract

**Background:** Wastewater monitoring is a valuable tool to track community-level disease trends. However, the extent to which vulnerable populations have been included in statewide wastewater monitoring programs remains unstudied.

**Objectives:** We conducted a geospatial analysis to examine (1) the representativeness of wastewater data collected through the North Carolina Wastewater Monitoring Network as of June 2022, and (2) the potential of wastewater data to generalize to unsewered populations in the county.

**Methods:** After intersecting wastewater treatment plant (WWTP) service areas (sewersheds) with census block and tract boundaries for 38 WWTPs across 18 counties, we compared the demographics and social vulnerability of (1) people residing in sewersheds of monitored WWTPs with countywide and statewide populations, and (2) people connected to any sewer system—regardless of inclusion in wastewater monitoring—with unsewered populations. We flagged differences greater than +/-5 percentage points or percent (for categorical and continuous variables, respectively) and noted which were statistically significant (i.e., greater than twice the margin of error).

**Results:** As a whole, populations in monitored sewersheds resembled the statewide population on most demographics analyzed, with a few exceptions. When multiple WWTPs were monitored within a county, their combined service populations resembled the countywide population, although populations in individually monitored sewersheds sometimes differed from the countywide population. In nine counties for which we had comprehensive sewershed maps, we found that sewered residents had higher social vulnerability, a greater share of Hispanics and African Americans, lower income, and lower educational attainment than unsewered residents.

**Discussion:** Our results suggest that wastewater monitoring in North Carolina well represents the larger community. Ongoing analyses will be needed as sites are added or removed. The approach we present here can be used to ensure that wastewater surveillance programs nationwide are implemented in a manner that informs equitable public health decision-making.

## Introduction

Early in the COVID-19 pandemic, clinical testing was restricted due to mass test kit shortages across the United States. Access to testing—a critical public health resource—aligned with structural disparities, with inequities among minority, uninsured, and rural groups (Rader et al. 2020). In poorer areas, there were fewer testing sites per person, and those sites were located farther away (Kim 2020). Communities that were majority Black and Hispanic were also more likely to face longer wait times and understaffed testing centers.

Recognizing that a better way existed to monitor population-wide infection levels, hundreds of communities launched wastewater testing for the SARS-CoV-2 virus that causes COVID-19. Wastewater monitoring can cover a much broader swath of the population than clinical testing, and taps into existing sanitation infrastructure, providing a practical and scalable solution to public health surveillance (Keshaviah 2021). In the United States, 16,000 wastewater treatment plants capture sewage from roughly 75% of the population (US EPA Office of Water, 2015). Worldwide, researchers estimate that roughly 1 in 4 people is connected to a wastewater treatment plant (Hart and Halden 2020). Critically, wastewater monitoring captures health biomarkers of sewered populations regardless of whether they visit a testing site or doctor, and regardless of whether they have symptoms, since people with asymptomatic infections also shed viral particles into their stools (Noh et al. 2020; Zhou et al. 2020).

Despite the potential of wastewater monitoring to improve health equity, resource constraints may inhibit equitable access to this innovative approach to public health surveillance. Prior to COVID-19, wastewater monitoring for diseases and controlled substances rarely occurred in low-or middle-income countries (LMICs). Of the fourteen countries that had routinely employed environmental surveillance for poliovirus as part of the Global Polio Eradication Initiative, ten (71%) were high-income countries (Hovi et al. 2012). Likewise, of the 37 member countries in the Sewage analysis CORe group—Europe network, which coordinates international wastewater studies on drug use in and beyond Europe, only 5 (14%) are LMICs (González-Mariño et al. 2020; World Bank n.d.). Even after the global expansion of wastewater testing to help officials worldwide manage the coronavirus pandemic, research has shown that monitoring has primarily occurred in high-income countries (Naughton et al. 2021).

While wastewater monitoring has the potential to overcome disparities in public health surveillance, little research has been conducted to determine the comparability of sewered and unsewered populations with respect to demographics and social vulnerability, and whether communities included in state and national wastewater monitoring programs resemble to those not being monitored. The National Academies Sciences, Engineering, and Medicine (2023) report, which stressed the importance of equity in national wastewater monitoring efforts, implied that because many unsewered households are in rural areas, and because rural areas tend to be more disadvantaged than urban areas, it follows that unsewered populations are more disadvantaged than sewered populations. However, an analysis of data from the 2019 American Household Survey found the opposite to be true–that septic households are more economically *advantaged* than sewered households–with the pattern upheld even when analyses were stratified by urbanicity (Olesen et al. 2022). Given that the Centers for Disease Control and Prevention’s National Wastewater Surveillance System (CDC NWSS) will continue to fund state, local, and tribal wastewater programs through at least 2025, officials need a way to ensure that wastewater sampling sites are selected in a manner that promotes health equity.

To assess how representative wastewater data collected during the COVID-19 pandemic are and could be, we conducted detailed geospatial analyses to answer two key questions: (1) Are sewered populations monitored through wastewater surveillance representative of the counties they come from with respect to demographics and social vulnerability? (2) How similar are the demographics and social vulnerability of communities that are and are not connected to a sewer system (regardless of inclusion in a wastewater monitoring program)? We focused our analysis on North Carolina, one of the first eight states funded by CDC NWSS.

## Methods

### Study site selection

Across both sets of analyses, we analyzed the service populations of 38 WWTPs in 18 counties, including 25 actively monitored WWTPs as of June 2022, 1 previously monitored WWTP, and 12 WWTPs not monitored by the NCWMN.

The North Carolina Wastewater Monitoring Network (NCWMN) collects wastewater samples from municipal wastewater treatment plants (WWTPs) across the state twice weekly and analyzes these samples to quantify SARS-CoV-2 viral concentrations. As of June 2022, the NCWMN routinely analyzed samples from 25 WWTPs across 17 of 100 NC counties, covering roughly one-fourth of the state’s population. Sites were added to the state’s wastewater monitoring program in stages. Because the initial group of sites came from a COVID-19 wastewater surveillance pilot project coordinated by universities (Noble et al. 2021), they were mainly located in urban centers near university labs that had the capacity analyze wastewater samples. Over time, the NCWMN expanded to include sites in other areas of the state, including the rural mountainous region in Western North Carolina, and underserved communities with higher social vulnerability, low COVID-19 vaccination rates, or both (NCDHHS 2021). In addition, five sites were added to the NCWMN when the Wake County health department initiated a local wastewater monitoring program (Supplemental Materials (SM) Figure S1).

To assess the representativeness of wastewater data, we conducted two sets of analyses. First, we analyzed the demographic and social vulnerability characteristics of people living in monitored sewersheds versus the general population, comparing: (A) individual monitored sewershed populations with their respective countywide population, (B) combined monitored sewershed populations, aggregated to the county level (in the case that multiple WWTPs were monitored within a county), with the respective countywide population and (C) combined monitored sewershed populations, aggregated to the state level, with the statewide population. Second, we compared the demographics and social vulnerability of sewered and unsewered populations in nine counties for which we could wholly identify the county’s sewered population using geospatial shapefiles delineating the service areas of all municipal WWTPs in the county with a treatment capacity of more than 0.5 million gallons per day. The nine counties covered rural and non-rural counties from across the state and included eight counties actively participating in NCWMN as of June 2022 plus Chatham County, which had previously participated in NCWMN. This latter analysis enabled us to evaluate the comparability of populations that can and cannot contribute to wastewater monitoring in the future.

### Community demographic and social vulnerability data

To summarize population demographics and social vulnerability for sewered and unsewered populations, we constructed 23 variables that represent five conceptual domains: demographics, health, housing and transportation, social vulnerability indices, and socio-economic status (SES) (SM Table S1). Most variables clearly fall within one of the five domains, while others straddle multiple domains. We grouped English proficiency within SES because language skills are often related to educational attainment and job prospects. Variables describing race and ethnicity came from the 2020 United States Census Redistricting Data, which were available at the block level geography. Since no other demographic variables were available in the 2020 Census data at the time of our analysis, we also analyzed variables from the 2015-2019 American Community Survey (2015-19 ACS) that captured age, gender, health insurance status, level of education, wealth, English proficiency, housing, employment, and disability status, all of which were available at the tract level geography. Lastly, we downloaded a shapefile with information on the CDC’s social vulnerability index (SVI), which were available at the tract level; we summarized the overall SVI percentile rank and the ranks for each of the four SVI themes (socioeconomic status, household composition and disability, minority status and language, and housing type and transportation).

### Geospatial analysis

To prepare the data for a geospatial analysis, we cleaned and joined tabular Census data to TIGER/Line tract or block polygons (US Census Bureau, 2022). We filtered the data to include only the counties in the study area described above. Polygon shapefiles for monitored sewersheds were shared by the NCWMN. For analyses of sewered versus unsewered populations, we merged shapefiles of NCWMN monitored sewersheds with shapefiles of WWTPs not monitored by the NCWMN, which we obtained from NC OneMap (North Carolina Department of Information Technology, 2022), to create a single county-level sewered polygon. We then created polygons that captured the county’s unsewered population by removing the sewered polygon from the county polygon.

To summarize the demographics, SES, and SVI of populations residing in the geographies of interest– including monitored individual sewersheds, monitored combined sewersheds, monitored counties, sewered county areas, unsewered county areas, and the state–we performed spatial intersections, selecting those census blocks or tracts that intersected each polygon of interest. We merged the selected tracts or blocks into a single polygon and calculated summary statistics: percentages that captures the proportion of the total population represented by different demographic groups, the average median household income, and population-weighted averages of SVI ranks. All analyses were performed using either ArcGIS Pro 2.9 (ESRI Inc. 2021) or R version 4.1.3 (R Core Team 2022) using the *sf* (Pebesma 2018) and *tidycensus* (Walker and Herman 2022) packages.

We designated potentially meaningful differences between populations using a threshold of +/-5 percentage points (pp) for categorical variables (demographic, SES variables, and SVI percentile ranks), and a threshold of +/-5 percent (%) for continuous variables (median household income). We chose this approach to be conservative and ensure that we did not overlook smaller disparities that were within reported margins of error (MOEs). This is especially relevant for a health-equity focused analysis because smaller groups often have larger MOEs, but a lack of statistical significance should not be interpreted as a lack of meaningful findings. For completeness, we also flagged whether such differences were statistically significant—i.e., whether the differences were within twice of the reported margins of error (MOEs)—for ACS 2015-2019 variables (note: 2020 Census data and SVI data did not include MOEs at the time of this analysis).

## Results

### Characteristics of WWTPs participating in the NC Wastewater Monitoring Network

The WWTPs participating in the NCWMN as of June 2022 covered a broad geographic area of the state and had service populations that ranged in size from 3,500 to 550,000 people and accounted for 1% (Raleigh 3) to 60% (City of Wilson) of the county’s population (Table 1). More detailed environmental meta data of the wastewater monitoring program can be found in SM Table S2 and a previous publication (Keshaviah et al. 2023). In three of the 17 counties analyzed, multiple WWTPs were being monitored, which together accounted for 33% (Mecklenburg), 54% (New Hanover), and 75% (Wake) of the respective county’s population.

**Table 1.** North Carolina sites monitored by the NCWMN as of June 2022.

| Site name | County name | Catchment<br>population size | County<br>population size | % of the county<br>population served |
| --- | --- | --- | --- | --- |
| Laurinburg | Scotland | 15,527 | 34,823 | 45% |
| Tuckaseegee | Jackson <sup>a</sup> | 13,296 | 43,109 | 31% |
| Marion | McDowell | 8,459 | 45,756 | 18% |
| Beaufort | Carteret <sup>a</sup> | 3,500 | 69,473 | 5% |
| Roanoke Rapids | Halifax | 14,320 | 69,493 | 21% |
| City of Wilson | Wilson | 49,384 | 81,801 | 60% |
| Chapel Hill – Carrboro | Orange | 78,141 | 148,476 | 53% |
| Greenville | Pitt <sup>a</sup> | 89,616 | 180,742 | 50% |
| Wilmington City | New Hanover <sup>a</sup> | 58,361 | 234,473 | 25% |
| New Hanover County (North) | New Hanover <sup>a</sup> | 67,743 | 234,473 | 29% |
| South Durham | Durham <sup>a</sup> | 108,105 | 321,488 | 34% |
| Fayetteville -Rockfish Creek | Cumberland | 151,589 | 335,509 | 45% |
| MSD of Buncombe County | Buncombe | 173,000 | 378,608 | 46% |
| Winston Salem - Salem | Forsyth <sup>a</sup> | 178,000 | 382,295 | 47% |
| Jacksonville | Onslow | 41,819 | 204,576 | 20% |
| Greensboro, North Buffalo | Guilford | 135,821 | 537,174 | 25% |
| Charlotte 1 | Mecklenburg <sup>a</sup> | 68,685 | 1,110,356 | 6% |
| Charlotte 2 | Mecklenburg <sup>a</sup> | 182,501 | 1,110,356 | 16% |
| Charlotte 3 | Mecklenburg <sup>a</sup> | 120,000 | 1,110,356 | 11% |
| Raleigh | Wake <sup>a</sup> | 550,000 | 1,111,761 | 49% |
| Raleigh 2 | Wake <sup>a</sup> | 37,020 | 1,111,761 | 3% |
| Raleigh 3 | Wake <sup>a</sup> | 7,648 | 1,111,761 | 1% |
| Cary 1 | Wake <sup>a</sup> | 84,189 | 1,111,761 | 8% |
| Cary 2 | Wake <sup>a</sup> | 74,331 | 1,111,761 | 7% |
| Cary 3 | Wake <sup>a</sup> | 75,886 | 1,111,761 | 7% |

Note: Sites are listed in order of ascending county population size. MSD=Metropolitan Sewerage District; WWTP=wastewater treatment plant.
<sup>a</sup> Indicates counties included in the sewered vs unsewered county analysis. Chatham county was not actively being monitored in June 2022 so it is not included here, but it is included in the sewered versus non-sewered analysis.

### Comparison of monitored sewershed with state and county populations

As a whole, populations residing in the 25 sewersheds monitored through NCWMN resembled the statewide resident population. Differences between the two groups amounted to less than +/-5 pp or 5% for 15 of the 23 variables analyzed, including: demographics (percent female, percent African American, percent Asian, percent American Indian/Alaska Native, percent Native Hawaiian or Pacific Islander, percent 65 years and older, percent Hispanic), health status (percent with disability, percent without health insurance), housing and transportation (percent of households without a vehicle, percent group quarters), housing and transportation vulnerability (based on the SVI), and SES (percent below federal poverty line, percent unemployed, percent limited English proficiency) (Figure 1A). However, monitored sewershed populations had fewer White residents (i.e., more minorities), lower overall social vulnerability but higher minority and language vulnerability, a higher number of houses with five or more units, greater educational attainment, a higher median household income and lower socioeconomic vulnerability, and lower vulnerability related to household composition and disability compared to the statewide population (Figure 1B). Among these potentially meaningful differences, only educational attainment reached statistical significance (SM Table S3).

**Figure 1.**
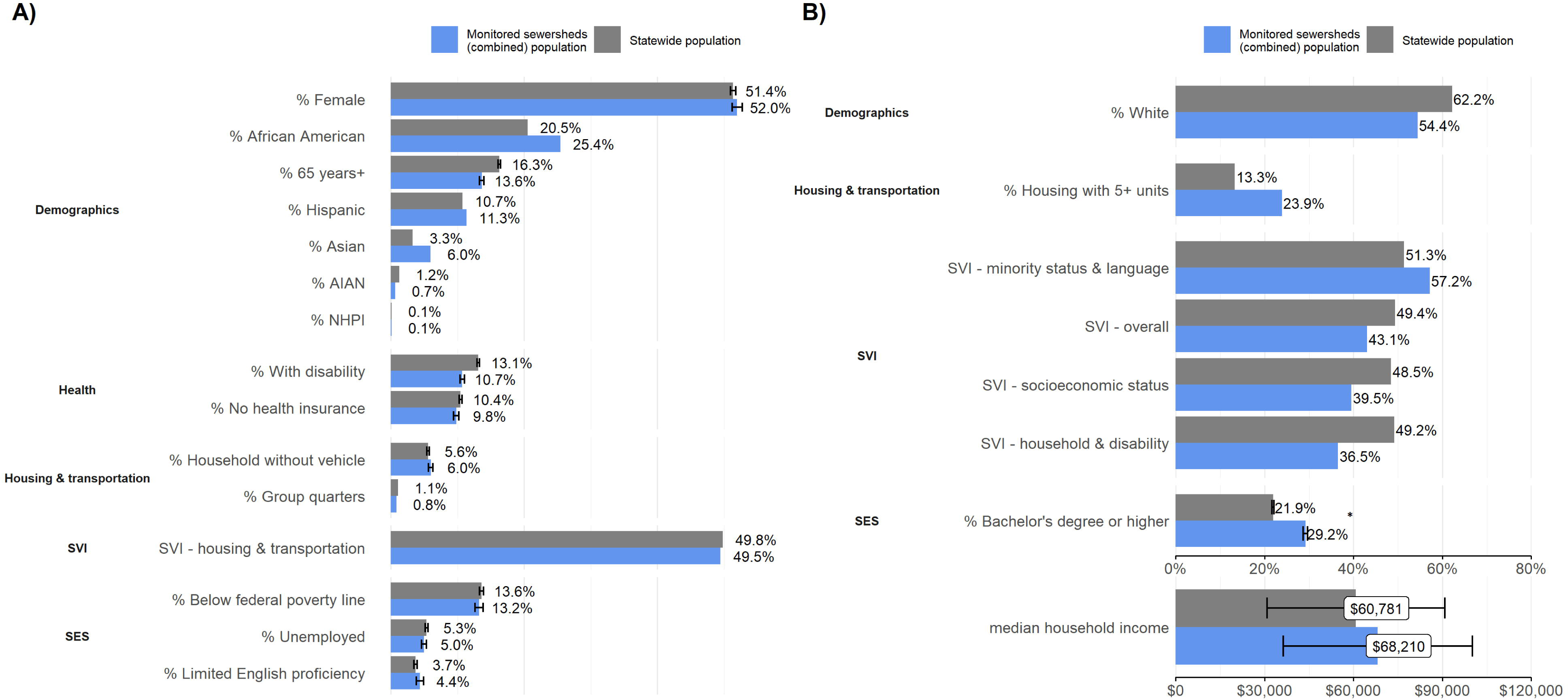
Demographic characteristics between the population in monitored sewersheds and the statewide population: A) difference between the population covered by monitored sewersheds and the statewide population is smaller than 5 percentage points; B) the difference between the population covered by monitored sewersheds and the statewide population equals or is greater than 5 percentage points. Note: The error bar represents the 95% confidence interval. 95% confidence interval which was only calculated for variables in the ACS 2015-2019 data because the MOE information wasn’t available for 2020 census at the time of the analysis. SVI is not a percent of the total population but a percentile rank. Only one variable (% Bachelor’s degree or higher) has a statistically significant difference (the difference is greater than twice the margin of error, shown with an asterisk (*)).

When we compared the populations living in the sewersheds monitored by the NCWMN with their respective countywide populations, we found that sewersheds had wide-ranging demographic and social characteristics. Combined monitored sewershed populations generally resembled their countywide population with respect to demographics (percent female, percent over 65 years old, percent Hispanic), health status (percent uninsured, percent disability), and SES (percent limited English speaking, percent below federal poverty, percent unemployed). However, there was a +/-5 pp or 5% difference between at least one combined monitored sewershed and the county for 12 out of the 23 variables, with the largest observed differences related to race, social vulnerability, median household income, and housing with greater than five units (Figure 2). Monitored sewershed populations had a lower share of Whites compared to countywide populations in 12 of 17 counties (shaded in blue), while African Americans made up a higher share of the monitored sewershed population in 14 of 17 counties, with the largest differences generally occurring in sewersheds in the eastern part of the state (shaded in red). In one county (Jackson County), we observed a potentially meaningful difference in the share of American Indian and Alaska Native residents, which was lower in the monitored sewershed than in the county (note: Jackson County borders the Qualla Boundary, which is home to the sovereign nation of the Eastern Band of the Cherokee Indians). Overall SVI ranks were similar between monitored sewershed populations and countywide populations. However, evaluating the SVI themes individually showed that either or both minority and language vulnerability and housing and transportation vulnerability were higher in most (15 out of 17) of monitored sewersheds than countywide (Figure 3). Lastly, the difference in median household income was wide-ranging across sewersheds (−19.8% to +5.8%), with nine sewershed populations having a higher median household than the county and eight having a lower median household income.

**Figure 2.**
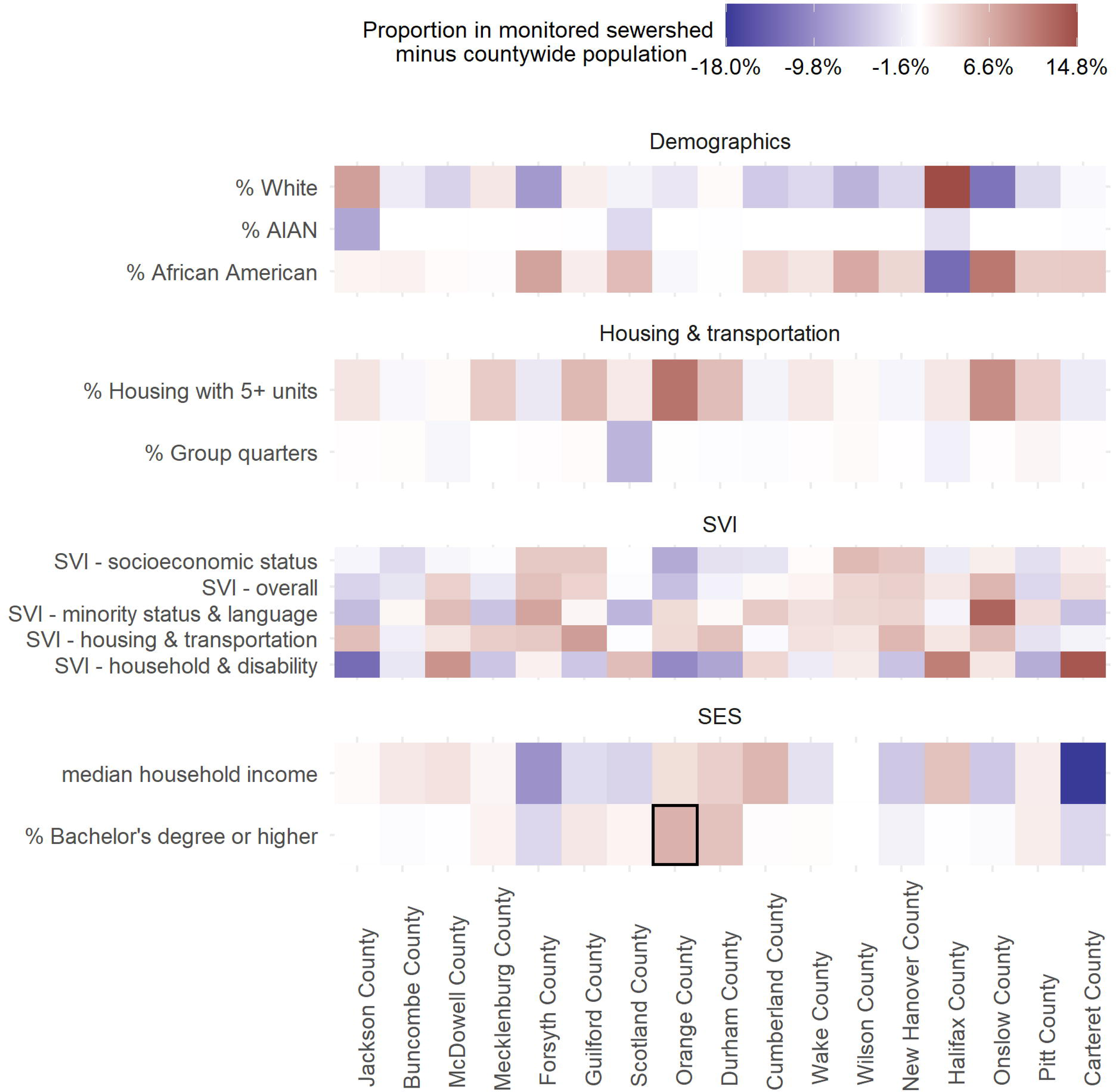
Demographic differences between the population in NCWMN monitored sewersheds and the respective counties. Note: Only demographic variables with more than a 5% difference (monitored – county) are included. For percentage variables, the difference is calculated by the monitored population minus the county population (i.e. percentage point difference). For income, the difference is calculated as the percent difference (i.e. income for the monitored population minus income for the county population, then divided by the income for the county population). Counties are ranked from west to east based on the location of the county centroids. Blocks highlighted with black outline are both meaningfully different and statistically significantly different (difference greater than twice the margin of error).

**Figure 3.**
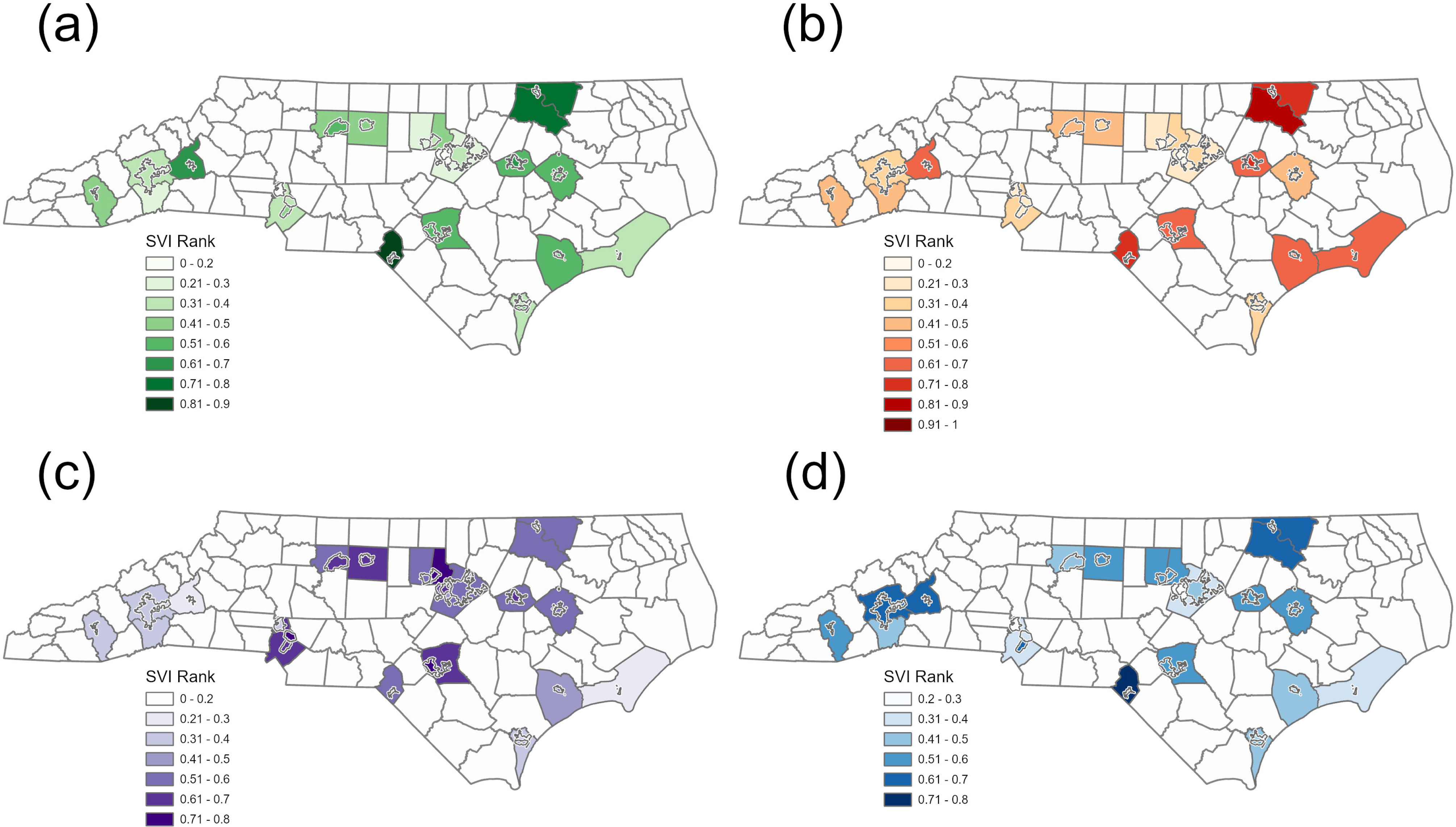
Social vulnerability percentile rankings in individual monitored NC sewersheds and respective counties. Maps show the four SVI themes: (a) socioeconomic status, (b) household composition and disability, (c) minority and language, and (d) housing and transportation.

In the three counties with multiple monitored WWTPs, we noted that populations in the individual monitored sewersheds had differing degrees of similarity to the countywide population. In all three counties, we observed meaningful differences in race, median household income, social vulnerability, educational attainment, and housing with 5 or more units that were not evident when the individual sewersheds were combined and analyzed as a single geographic unit (SM Table S5). For example, in Wake County, the combined sewershed SVI rankings resembled the county SVI rankings even though the six individual sewersheds showed a wide range of SVI rankings across all four SVI themes: socioeconomic status (individual ranged from 0.12-0.51, combined=0.27, county=0.27), household composition and disability (individual=0.16-0.74, combined=0.28, county=0.29), minority status and language (individual=0.44-0.76, combined=0.59, county=0.56), and housing and transportation (individual=0.25-0.63, combined=0.42, county=0.40) (Figure 3). Notably, residents in two Wake County sewersheds, Raleigh and Raleigh 3, appeared to be more disadvantaged than other Wake sewersheds and countywide residents, given their higher social vulnerability overall and across all themes, coupled with a lower educational achievement and lower median household income.

### Comparison of sewered and unsewered populations

When we compared the characteristics of sewered and unsewered populations in nine counties with complete information on WWTP service populations, we found that only four of 23 variables did not meaningfully differ—including percent Asian, percent Native Hawaiian and Pacific Islander, percent female, and percent unemployed—while the remaining 19 variables differed in at least one county (Figure 4). Most notably, we found differences in racial and ethnic makeup, median household income, and social vulnerability. In most of the 9 counties, Hispanics and African Americans made up a greater share of the sewered population than the unsewered population (shaded in red). However, the magnitude and direction of effect sometimes varied across counties with a −4.6 to 28.0 pp difference in the share of Hispanics and a 2.4 to 18.0 pp difference in the share of African Americans. We also found that in all but one county (Pitt County), the median household income was lower in the sewered population than the unsewered population, with differences ranging from −30.0% to −0.2%. Contrary to our finding that population residing in NCWMN monitored sewersheds have higher educational attainment than the statewide population, we saw that educational attainment was lower in the sewered population than the unsewered population in 7 of 9 counties (all but Forsyth County and Pitt County), ranging from a −17.0 to −0.2 pp difference. This difference was also statistically significant for Durham and Chatham counties (SM Table S4). Finally, in 7 of 9 counties (all but Jackson County and Pitt County), we found that overall social vulnerability and vulnerability based on each of the four SVI themes were higher among the sewered population than the unsewered population.

**Figure 4.**
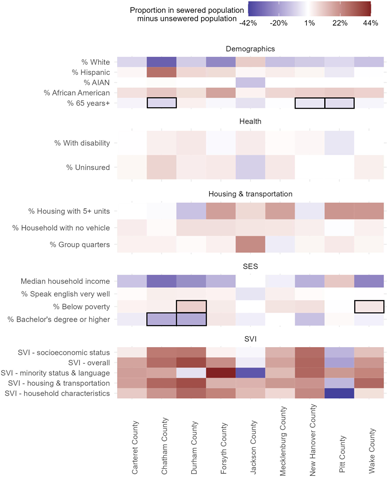
Demographic differences between the sewered population and the unsewered population. Note: Only demographic variables with more than a 5% difference (sewered – unsewered) are included. For percentage variables, the difference is calculated by the sewered population minus the unsewered population (i.e. percentage point difference). For income, the difference is calculated as the percent difference (i.e. income for the sewered population minus income for the unsewered population, then divided by the income for the unsewered population). Counties are ranked from west to east based on the location of the county centroids. Blocks highlighted with black outline are both meaningfully different and statistically significantly different (difference greater than twice the margin of error).

## Discussion

Our findings indicate that residents of 25 WWTP sewersheds monitored by NCMWN as of June 2022 represent the broader North Carolina population fairly well. The share of elderly residents was comparable between monitored sewersheds and state- and county-wide populations, suggesting that wastewater data is capturing infections among groups that experienced more severe COVID-19 disease and higher mortality rates. Further, the share of residents with health insurance, English proficiency, and a vehicle was similar between residents of monitored sewersheds and the respective countywide population, suggesting that there is little bias in the data related to health care access. Given the similarities of demographic and social vulnerability dimensions across most counties, wastewater data collected through the NCWMN can be validly used to describe state and county population health.

We did note a few differences that have implications related to health equity. First, we noted that educational attainment was significantly higher among the monitored population than statewide. This finding is important because lower educational attainment has been shown to associate with lower receptivity to public health messaging and higher vaccine hesitancy (Anakpo and Mishi 2022). This may be explained by the fact that the initial sites chosen for monitoring were located near the university laboratories that piloted wastewater testing. Second, in the three counties with multiple monitored sewersheds, differences between characteristics of residents in individual monitored sewersheds versus countywide suggest that, had NCWMN monitored only one of the sewersheds in the county, or if the site composition in these counties changes over time, the wastewater data may not be representative of the larger population. Third, we found that sewered populations have significantly lower educational attainment, a lower share of elderly residents, and a higher share living below the poverty level compared to unsewered residents. In other words, sewered populations may be more vulnerable than unsewered populations, suggesting that wastewater data is likely to capture the health status of vulnerable residents.

Our analyses also highlight that minority populations may be over-represented in the state’s wastewater data. African Americans represented a higher share of monitored sewershed residents than countywide, while Whites often comprised a smaller share of monitored sewershed residents than county-or state-wide. Also, vulnerability related to minority status and language was greater in monitored sewersheds than statewide. Together, these findings have two implications. First, they suggest that, by better representing potentially vulnerable racial and ethnic minorities, wastewater monitoring may have filled critical gaps in clinical case data, which likely underrepresented Black and Hispanic communities early in the pandemic. More recent research has also found that, in the summer of 2022 (when the Omicron variant was dominant), COVID-19 cases were severely underestimated, with differences particularly pronounced among Black and Hispanic populations, younger adults ages 18 to 24, and those with lower income and less education (Qasmieh et al. 2023). The second implication of having a higher share of minority residents in monitored sewersheds versus county-or state-wide is that it creates a risk of inaccurate health messaging to the public. Given that racial and ethnic minorities have seen higher SARS-CoV-2 infection rates than White, non-Hispanic populations (CDC 2023; Mackey et al. 2021), wastewater data that overrepresents these groups could lead to inflated COVID-19 infection estimates.

Although we found that populations monitored by NCWMN were generally representative of the larger county- and state-wide population, there were some limitations to our analysis. First, our findings may not hold as monitoring sites are added or removed in the future. At the time of our analysis, which was based on a snapshot as of June 2022, the NCWMN was monitoring 25 sewersheds in 17 counties. Roughly one year later— as of June 14, 2023—the NCWMN monitors 50 sites in 32 counties. Accordingly, continued efforts will be needed to uncover any bias that emerges in the wastewater data. Expanding statewide monitoring to include community onsite wastewater treatment systems could also strengthen equitable coverage. This strategy may be particularly useful in NC, since roughly 50% of state residents use septic systems (NCDHHS 2022), and we found that NC’s sewered and unsewered populations differ with respect to demographics and social vulnerability.

A second limitation is that our geospatial analysis may have misclassified some residents as being part of the monitored sewershed population when they were not. To aggregate data to the sewershed level, we utilized a spatial intersect to select tracts or blocks that intersected the sewershed polygon. This selection method ensured that the selection for small or narrow sewersheds was greater than zero, but since neither geography perfectly aligns with the sewershed boundary, it possibly over-estimated the number of persons in the sewershed when tracts or blocks partially extend outside the sewershed boundary. Future studies using hi-resolution gridded population data would more accurately assign populations to the sewershed (Depsky et al. 2022). We also assumed that all homes inside the sewershed boundary are connected to the sewer, even though some may use onsite septic systems. We were unable to discern the magnitude or direction of the resulting bias from this assumption because septic system location data are not readily available for the state.

A third limitation is that our analysis could not examine representativeness related to SARS-CoV-2 fecal shedding rates. People that shed little or no virus in their feces will not be represented in wastewater data, and preliminary research suggests that demographic and geographic features may influence viral shedding rates. For example, early in the pandemic, Parasa et al. (2020) found that fecal shedding rates varied substantially across eight studies included in their meta-analysis, estimating that, on average, 41% of confirmed COVID-19 cases (range=17% to 80%) shed the virus in their stools. More recently, Prasek et al. (2023) noted differences in estimated shedding rates across communities of differing ages, ethnicity, and socioeconomic composition, as well as over time, as the dominant variant changed (though it is worth noting that these findings were subject to ecological fallacy and lacked the use of multivariate regression modeling to control for confounding factors).

Lastly, it is important to interpret our findings in the context of known limitations and biases in the underlying Census data. Data on race and ethnicity collected during the 2020 US Census were subjected to a new disclosure avoidance system called differential privacy, which added statistical noise to the published data products to shield sensitive information from discovery. However, the amount of statistical noise added to the data is not constant and small demographic groups and geographic areas were infused with more noise to reduce the risk of re-identification (Garfinkel 2022). We aggregated Census block data to larger geographies, which should minimize inaccuracies associated with differential privacy. Moreover, the Demographic Analysis, one of the leading indicators of data quality for decennial censuses, showed a record undercount of Hispanics during the 2020 Census (Cohn and Passel 2022). Although we did not discover a meaningful difference in the percent of the population that was Hispanic between monitored sewershed and the county or the state, it is possible that a difference was obscured by undercounting.

## Conclusions

Evidence-based public health decisions need to be informed by complete, high-quality data that represent the community. Our analyses confirm that wastewater data from across North Carolina well represents county- and state-wide populations, and that by capturing the health information of vulnerable populations better than clinical data—particularly when clinical resources are in short supply— wastewater data can promote health equity. The in-depth geospatial analyses we conducted here identify underlying bias in wastewater data, and in doing so, can also help officials recognize when and how to tailor public health messaging and response accordingly.

## Data Availability

The data that support the findings of this study are available from the U.S. Census Data API https://www.census.gov/data/developers/data-sets/census-microdata-api.html. Shapefiles of the WWTP sewer service areas (not including those provided by NCWMN) are available from NC OneMap https://www.nconemap.gov/datasets/nconemap::type-a-current-public-sewer-systems-2004/about.

## Acknowledgements

The authors would like to acknowledge the contributions of several partners to this work, including Virginia Guidry, Ariel Christensen, and Steven Berkowitz from the North Carolina Department of Health and Human Services. Financial support for this research was provided by the authors’ institutions— Mathematica, NCDHHS, and North Carolina State University.

## Notes

### Competing Interest Statement

The authors have declared no competing interest.

### Funding Statement

Financial support for this research was provided by the authors institutions: Mathematica, North Carolina Department of Health and Human Services, and North Carolina State University

### Author Declarations

The study used ONLY aggregate human data from the American Community Survey and the publicly available NC COVID dashboard, which meets NCDHHS Division of Public Health data suppression guidelines.

## References

Anakpo G, Mishi S. 2022. Hesitancy of COVID-19 vaccines: Rapid systematic review of the measurement, predictors, and preventive strategies. Human Vaccines & Immunotherapeutics 18:2074716; doi:10.1080/21645515.2022.2074716.

CDC. 2023. Cases, Data, and Surveillance. Centers for Disease Control and Prevention. Available: https://www.cdc.gov/coronavirus/2019-ncov/covid-data/investigations-discovery/hospitalization-death-by-race-ethnicity.html [accessed 26 April 2023].

Cohn D, Passel JS. 2022. Key facts about the quality of the 2020 census. Pew Research Center.

Depsky NJ, Cushing L, Morello-Frosch R. 2022. High-resolution gridded estimates of population sociodemographics from the 2020 census in California. PLOS ONE 17:e0270746; doi:10.1371/journal.pone.0270746.

ESRI Inc. 2021. ArcGIS Pro (Version 2.9).

Garfinkel S. 2022. Differential Privacy and the 2020 US Census · Winter 2022. Available: https://mit-serc.pubpub.org/pub/differential-privacy-2020-us-census/release/1 [accessed 26 April 2023].

González-Mariño I, Baz-Lomba JA, Alygizakis NA, Andrés-Costa MJ, Bade R, Bannwarth A, et al. 2020. Spatio-temporal assessment of illicit drug use at large scale: evidence from 7 years of international wastewater monitoring. Addiction 115:109–120; doi:10.1111/add.14767.

Hart OE, Halden RU. 2020. Computational analysis of SARS-CoV-2/COVID-19 surveillance by wastewater-based epidemiology locally and globally: Feasibility, economy, opportunities and challenges. Science of The Total Environment 730:138875; doi:10.1016/j.scitotenv.2020.138875.

Hovi T, Shulman LM, van der Avoort H, Deshpande J, Roivainen M, De Gourville EM. 2012. Role of environmental poliovirus surveillance in global polio eradication and beyond. Epidemiol Infect 140:1–13; doi:10.1017/S095026881000316X.

Keshaviah A. 2021. Next steps for wastewater testing to help end this pandemic — and prevent the next one. STAT. Available at https://www.statnews.com/2021/06/24/wastewater-testing-infrastructure-improvements-public-health/

Keshaviah A, Huff I, Hu XC, Guidry V, Christensen A, Berkowitz S, et al. 2023. Separating signal from noise in wastewater data: An algorithm to identify community-level COVID-19 surges in real time. Proceedings of the National Academy of Sciences 120:e2216021120; doi:10.1073/pnas.2216021120.

Kim SR. 2020. Which Cities Have The Biggest Racial Gaps In COVID-19 Testing Access? FiveThirtyEight.

Mackey K, Ayers CK, Kondo KK, Saha S, Advani SM, Young S, et al. 2021. Racial and Ethnic Disparities in COVID-19–Related Infections, Hospitalizations, and Deaths. Ann Intern Med 174:362–373; doi:10.7326/M20-6306.

National Academies Sciences Engineering and Medicine,. 2023. Wastewater-based Disease Surveillance for Public Health Action |The National Academies Press. Available: https://nap.nationalacademies.org/catalog/26767/wastewater-based-disease-surveillance-for-public-health-action [accessed 7 April 2023].

Naughton CC, Roman FA, Alvarado AGF, Tariqi AQ, Deeming MA, Bibby K, et al. 2021. Show us the Data: Global COVID-19 Wastewater Monitoring Efforts, Equity, and Gaps. 2021.03.14.21253564; doi:10.1101/2021.03.14.21253564.

NCDHHS. 2022. EHS: On-Site Wastewater Treatment and Dispersal Systems Program Resources. Available: https://ehs.dph.ncdhhs.gov/oswp/resources.htm [accessed 26 April 2023].

NCDHHS. 2021. NCDHHS Doubles Wastewater Monitoring Sites to Track the Spread of COVID-19. Available: https://www.ncdhhs.gov/news/press-releases/2021/07/22/ncdhhs-doubles-wastewater-monitoring-sites-track-spread-covid-19 [accessed 27 January 2023].

Noble R, de los Reyes F, Harris A, Stewart J, Cahoon L, Engel L, et al. 2021. Tracking SARS-CoV-2 in the Wastewater Across a Range of North Carolina Municipalities.

Noh JY, Yoon JG, Seong H, Choi WS, Sohn JW, Cheong HJ, et al. 2020. Asymptomatic infection and atypical manifestations of COVID-19: Comparison of viral shedding duration. Journal of Infection 81:816–846; doi:10.1016/j.jinf.2020.05.035.

North Carolina Department of Information Technology,. 2022. NC OneMap. Available: https://www.nconemap.gov/ [accessed 7 April 2023].

Olesen SW, Young C, Duvallet C. 2022. The Effect of Septic Systems on Wastewater-Based Epidemiology.

Parasa S, Desai M, Thoguluva Chandrasekar V, Patel HK, Kennedy KF, Roesch T, et al. 2020. Prevalence of Gastrointestinal Symptoms and Fecal Viral Shedding in Patients With Coronavirus Disease 2019: A Systematic Review and Meta-analysis. JAMA Network Open 3:e2011335; doi:10.1001/jamanetworkopen.2020.11335.

Pebesma E. 2018. Simple Features for R: Standardized Support for Spatial Vector Data. The R Journal 10:439–446; doi:10.32614/RJ-2018-009.

Prasek SM, Pepper IL, Innes GK, Slinski S, Betancourt WQ, Foster AR, et al. 2023. Variant-specific SARS-CoV-2 shedding rates in wastewater. Science of The Total Environment 857:159165; doi:10.1016/j.scitotenv.2022.159165.

Qasmieh SA, Robertson MM, Teasdale CA, Kulkarni SG, Jones HE, McNairy M, et al. 2023. The prevalence of SARS-CoV-2 infection and long COVID in U.S. adults during the BA.4/BA.5 surge, June–July 2022. Preventive Medicine 169:107461; doi:10.1016/j.ypmed.2023.107461.

R Core Team. 2022. R: A Language and Environment for Statistical Computing. R Foundation for Statistical Computing:Vienna, Austria.

Rader B, Astley CM, Sy KTL, Sewalk K, Hswen Y, Brownstein JS, et al. 2020. Geographic access to United States SARS-CoV-2 testing sites highlights healthcare disparities and may bias transmission estimates. J Travel Med 27:taaa076; doi:10.1093/jtm/taaa076.

US Census Bureau,. 2022. TIGER/Line Shapefiles. Census.gov. Available: https://www.census.gov/geographies/mapping-files/time-series/geo/tiger-line-file.html [accessed 7 April 2023].

US EPA Office of Water,. 2015. Learn about Small Wastewater Systems. Available: https://www.epa.gov/small-and-rural-wastewater-systems/learn-about-small-wastewater-systems [accessed 27 January 2023].

Walker K, Herman M. 2022. tidycensus: Load US Census Boundary and Attribute Data as “tidyverse” and ‘sf’-Ready Data Frames.

World Bank. n.d. World Bank Country and Lending Groups – World Bank Data Help Desk. Available: https://datahelpdesk.worldbank.org/knowledgebase/articles/906519-world-bank-country-and-lending-groups [accessed 7 December 2022].

Zhou R, Li F, Chen F, Liu H, Zheng J, Lei C, et al. 2020. Viral dynamics in asymptomatic patients with COVID-19. International Journal of Infectious Diseases 96:288–290; doi:10.1016/j.ijid.2020.05.030.429

