## Supplemental Material for "Assessing health equity in wastewater monitoring programs: Differences in the demographics and social vulnerability of sewered and unsewered populations across North Carolina"

Table S1 Demographic variables included in this analysis

| **Group** | **Variable Description** | **Variable ID** | **Source** | **Geography** |
| --- | --- | --- | --- | --- |
| Demographics | Percent 65+ years | S0101_C01_030 | ACS 2015-19 | tract |
|  | Percent female | B01001_026/B01001_001 | ACS 2015-19 | tract |
|  | Percent White | P1_003N/P1_001N | US Census 2020 | block |
|  | Percent African American | P1_004N/P1_001N | US Census 2020 | block |
|  | Percent Asian | P1_006N/P1_001N | US Census 2020 | block |
|  | Percent American Indian or Alaska Native | P1_005N/P1_001N | US Census 2020 | block |
|  | Percent Native Hawaiian or Pacific Islander | P1_007N/P1_001N | US Census 2020 | block |
|  | Percent Hispanic | P2_002N/P1_001N | US Census 2020 | block |
| Health | Percent with disability | DP02_0072P/B01001_001 | ACS 2015-19 | tract |
|  | Percent uninsured | S2701_C05_001/B01001_001 | ACS 2015-19 | tract |
| Housing and transportation | Percent households without a vehicle | DP04_0058P | ACS 2015-19 | tract |
|  | Percent of housing structures with 5 or more units | DP04_0011P + DP04_0012P + DP04_0013P | ACS 2015-19 | tract |
|  | Percent population group quarters | P5_001N | US Census 2020 | block |
| SVI | SVI - Overall vulnerability | RPL_THEMES | CDC SVI 2018 | tract |
|  | SVI - Socioeconomic status | RPL_THEME1 | CDC SVI 2018 | tract |
|  | SVI - Household composition & disability | RPL_THEME2 | CDC SVI 2018 | tract |
|  | SVI - Minority status & language | RPL_THEME3 | CDC SVI 2018 | tract |
|  | SVI - Housing type & transportation | RPL_THEME4 | CDC SVI 2018 | tract |
| Socioeconomic (SES) | Percent population with college or higher (25 yrs+) | (B06009_004 + B06009_005 + B06009_006)/B01001_001 | ACS 2015-19 | tract |
|  | Median household income | B19013_001 | ACS 2015-19 | tract |
|  | Percent below poverty | B17001_002/ B01001_001 | ACS 2015-19 | tract |
|  | Percent unemployed | DP03_0005/B01001_001 | ACS 2015-19 | tract |
|  | Percent population speaking English less than “well” | (B16005_007 + B16005_008 + B16005_012 + B16005_013 + B16005_017 + B16005_018 + B16005_022 + B16005_023 + B16005_029 + B16005_030 + B16005_034 + B16005_035 + B16005_039 + B16005_040 + B16005_044 + B16005_045) / B01001_001 | ACS 2015-19 | tract |

Table S2 Meta-information on wastewater infrastructure, sampling, and testing methods

| Site name | Capacity (MGD) | Mean  daily flow (MGD) | Sample type | Sampling start time | Sample volume (mL) | Median recovery efficiency (%) |
| --- | --- | --- | --- | --- | --- | --- |
| Laurinburg | 4 | 2.3 | Flow | 8:30am | 40 | 35 |
| Tuckaseigee | 3.5 | 1.04 | Time | 8:00am | 25 | 27 |
| Marion | 3 | 0.87 | Flow | 10:00am | 40 | 30 |
| Beaufort | 1.5 | 0.91 | Flow | 10:00am | 40 | 30 |
| Roanoke Rapids | 8.3 | 2.8 | Flow | 8:30am | 40 | 35 |
| City of Wilson | 14 | 8.2 | Flow | 6:30am | 40 | 34 |
| Chapel Hill - Carrboro | 14.5 | 7.5 | Flow | 7:00am | 40 | 32 |
| Greenville | 17.5 | 11.2 | Flow | 9:00am | 40 | 28 |
| Wilmington City (North) | 5.5 | 3.7 | Flow | 7:00am | 40 | 36 |
| New Hanover County (North) | 10.5 | 6.8 | Flow | 7:00am | 40 | 30 |
| South Durham | 20 | 8.4 | Flow | 6:45am | 40 | 29 |
| Fayetteville - Rockfish | 21 | 14.9 | Flow | 4:30am | 40 | 32 |
| MSD of Buncombe County | 40 | 21 | Flow | 9:00am | 40 | 30 |
| Winston Salem - Salem | 15.3 | 10.6 | Flow | 5:30am | 40 | 33 |
| Greensboro - North Buffalo | 18 | 13.7 | Flow | 8:00am | 40 | 38 |
| Charlotte 1 | 12 | 4.2 | Flow | 7:30am | 40 | 33 |
| Charlotte 2 | 28 | 17 | Flow | 8:00am | 40 | 38 |
| Charlotte 3 | 12 | 9.9 | Flow | 7:00am | 40 | 44 |
| Raleigh | 75 | 49.5 | Flow | 7:00am | 40 | 34 |
| Raleigh 2 | 3 | 2.5 | Flow | 8:00am | 40 | 26 |
| Raleigh 3 | 2.2 | 0.8 | Flow | 7:00am | 40 | 34 |
| Cary 1 | 12 | 6.9 | Flow | 6:30am | 40 | 26.5 |
| Cary 2 | 12.8 | 5.4 | Flow | 8:30am | 40 | 30 |
| Cary 3 | 18 | 6.5 | Flow | 7:00am | 40 | 41.5 |
| Jacksonville | 9 | 3.7 | Flow | 8:00am | 40 | 25 |

Note: The table shows meta-variables that substantially differed across sites. Wastewater sample analysis was conducted by the following three labs, Tuckaseigee by University of Wisconsin-Milwaukee, Raleigh 2, Raleigh 3, Cary 1, Cary 2, and Cary 3 by North Carolina State University, and the rest by University of North Carolina-Chapel Hill. Sampling generally occurred twice weekly, though was often less frequent around holidays, and occurred only weekly in Tuckaseigee before August 2021. The concentration method used was membrane filtration with MgCl_­­2_ (all sites) and acidification (all sites except Tuckaseigee). Extraction used the NUCLISENSE manual magnetic bead extraction kit (all sites except Tuckaseigee) or bead bashed HA filters on a KingFisher Flex system 96 well plates (Tuckaseigee only). All sites shared the following features: Sample location type = wastewater treatment plant, System type = separated, Sample mix = raw wastewater, pre-concentration storage temp = 4°C, PCR type = Digital droplet polymerase chain reaction (ddPCR), SARS-CoV-2 targets = N1 and N2, recovery control name = Bovine coronavirus (BCoV) vaccine, endogenous control = Pepper mild mottle virus (PMMoV), extraction blanks = yes. For additional details on sample analysis methods, see previous publications (1, 2). Sites are ordered by ascending county population size.

Flow = 24-hr flow-weighted composite; MGD = Million gallons per day; MSD = Metropolitan Sewerage District; Time = 24-hr time-weighted composite.

Table S3 Demographic characteristics between the population in monitored sewersheds and the countywide/statewide population

Attached TableS3.xlsx

Table S4 Difference in the demographic characteristics between the sewered and unsewered population

Attached TableS4.xlsx

Table S5 Differences in demographic characteristics between the population in individually monitored sewersheds or combined monitored sewersheds and the county population

A) **Demographics**

| **Site name** | **White** | **African American** | **Asian** | **Hispanic** | **65 years+** |
| --- | --- | --- | --- | --- | --- |
| Cary 1 | -5.7 | -9.7 | 18.8 | -3.9 | -2.5 * |
| Cary 2 | 13.3 | -11.4 | -1.5 | -0.8 | 4.1 * |
| Cary 3 | -5.8 | -11.0 | 21.8 | -5.3 | -1.1 |
| Raleigh | -4.9 | 7.6 | -4.0 | 1.7 | 0.3 |
| Raleigh 2 | 10.0 | -2.1 | -4.8 | -3.6 | -0.6 |
| Raleigh 3 | -18.4 | 21.6 | -7.6 | 4.8 | 5.2 |
| Combined Wake County sites (6) | -3.3 | 2.1 | 1.2 | 0.1 | 0.3 |
| Charlotte 1 | 29.2 | -20.3 | -3.2 | -6.4 | 3.9 * |
| Charlotte 2 | 5.3 | -4.0 | -2.5 | 2.1 | -0.5 |
| Charlotte 3 | -19.2 | 18.0 | 2.5 | -1.3 | -3.0 * |
| Combined Mecklenburg County sites (3) | 2.0 | 0.3 | -0.8 | -1.3 | -0.5 |
| New Hanover County, North | 2.6 | -2.4 | -0.1 | 0.2 | 1.0 |
| Wilmington City, North | -7.5 | 7.3 | -0.1 | 0.4 | -0.7 |
| Combined New Hanover County sites (2) | -3.5 | 3.3 | 0.0 | 0.3 | -0.5 |

B) **Social Vulnerability Index (SVI)**

| **Site name** | **Socioeconomic status** | **Household composition & disability** | **Minority status & language** | **Housing & transportat-ion** | **Overall SVI** |
| --- | --- | --- | --- | --- | --- |
| Cary 1 | -14.7 | -13.8 | 11.7 | -10.4 | -12.4 |
| Cary 2 | -10.2 | -9.9 | -9.1 | -14.9 | -12.7 |
| Cary 3 | -14.7 | -5.0 | 4.9 | -12.3 | -11.3 |
| Raleigh | 6.0 | 1.1 | 3.2 | 8.3 | 6.2 |
| Raleigh 2 | -7.1 | 6.5 | -12.4 | -7.2 | -7.0 |
| Raleigh 3 | 24.3 | 44.4 | 19.5 | 23.2 | 34.6 |
| Combined Wake County sites (6) | 0.4 | -1.8 | 2.6 | 2.4 | 1.0 |
| Charlotte 1 | -24.0 | -9.4 | -34.6 | -16.9 | -25.7 |
| Charlotte 2 | 11.3 | -0.7 | 0.4 | 23.0 | 12.7 |
| Charlotte 3 | 3.3 | -5.7 | 14.2 | -2.1 | -0.2 |
| Combined Mecklenburg County sites (3) | -0.4 | -5.0 | -5.2 | 4.1 | -2.0 |
| New Hanover County, North | -6.2 | -3.9 | 3.6 | -1.0 | -2.5 |
| Wilmington City, North | 12.1 | -6.0 | 10.9 | 12.2 | 10.9 |
| Combined New Hanover County sites (2) | 4.6 | -5.2 | 3.5 | 5.9 | 3.8 |

C) **Socioeconomic status (SES), health, and housing & transportation**

| **Site name** | **Bachelor's degree or higher** | **Median household income** | **Below federal poverty level** | **Unemployed** | **Limited English proficiency** | **No health insurance** | **Housing w/ 5+ units** |
| --- | --- | --- | --- | --- | --- | --- | --- |
| Cary 1 | 10.4 * | 17.9 | -3.5 * | -0.9 | 2.2 | -3.2 * | 7.7 |
| Cary 2 | 7.1 * | 12.8 | -3.4 * | -1.3 | -0.4 | -0.9 | -2.9 |
| Cary 3 | 9.6 * | 26.9 | -2.1 | 0.0 | 1.3 | -3.4 * | -8.0 |
| Raleigh | -2.5 * | -13.2 | 1.7 * | 0.3 | 0.1 | 1.1 * | 4.3 |
| Raleigh 2 | -2.7 | 3.1 | -2.1 | 1.7 | -2.4 | -2.0 | -5.4 |
| Raleigh 3 | -18.2 * | -28.7 | 2.1 | 5.1 | 5.5 | 13.4 | -18.7 |
| Combined Wake County sites (6) | 0.3 | -2.6 | -0.2 | 0.1 | 0.2 | 0.2 | 1.9 |
| Charlotte 1 | 4.4 * | 25.5 | -5.3 * | -0.9 | -5.2 * | -6.0 * | -10.9 |
| Charlotte 2 | 2.0 | -5.6 | 5.6 * | -0.1 | 2.6 * | 2.9 * | 12.8 |
| Charlotte 3 | -4.4 * | -18.0 | 0.6 | -0.7 | 0.5 | 0.7 | 3.1 |
| Combined Mecklenburg County sites (3) | 1.0 | 0.8 | 0.6 | -0.6 | -0.4 | -0.5 | 4.2 |
| New Hanover County, North | 0.5 | 11.6 | -2.8 * | 0.5 | -0.2 | -1.7 | -7.4 |
| Wilmington City, North | -1.0 | -10.4 | 5.3 * | -0.2 | 0.2 | -0.3 | 4.2 |
| Combined New Hanover County sites (2) | -1.2 | -4.9 | -0.3 | 0.6 | 0.1 | -0.6 | -0.9 |

Note: Only variables with more than a 5 percent (%) or percentage point (pp) difference (monitored sewershed - county) in at least one sewershed are included. For categorical variables (demographics, SVI, SES variables, health, and housing & transportation), the difference is calculated by the monitored sewershed population minus the county population (i.e. pp difference). For the continuous variable (median household income), the difference is calculated as income for the monitored sewershed population minus income for the county population, then divided by the income for the county population (i.e. % difference). Variables which have a statistically significant difference (the difference is greater than twice the margin of error (MOE)) are shown with an asterisk (*). MOE information was only available for variables in the ACS 2015-2019 data.

Figure S1 North Carolina map of sites included in the geospatial analysis

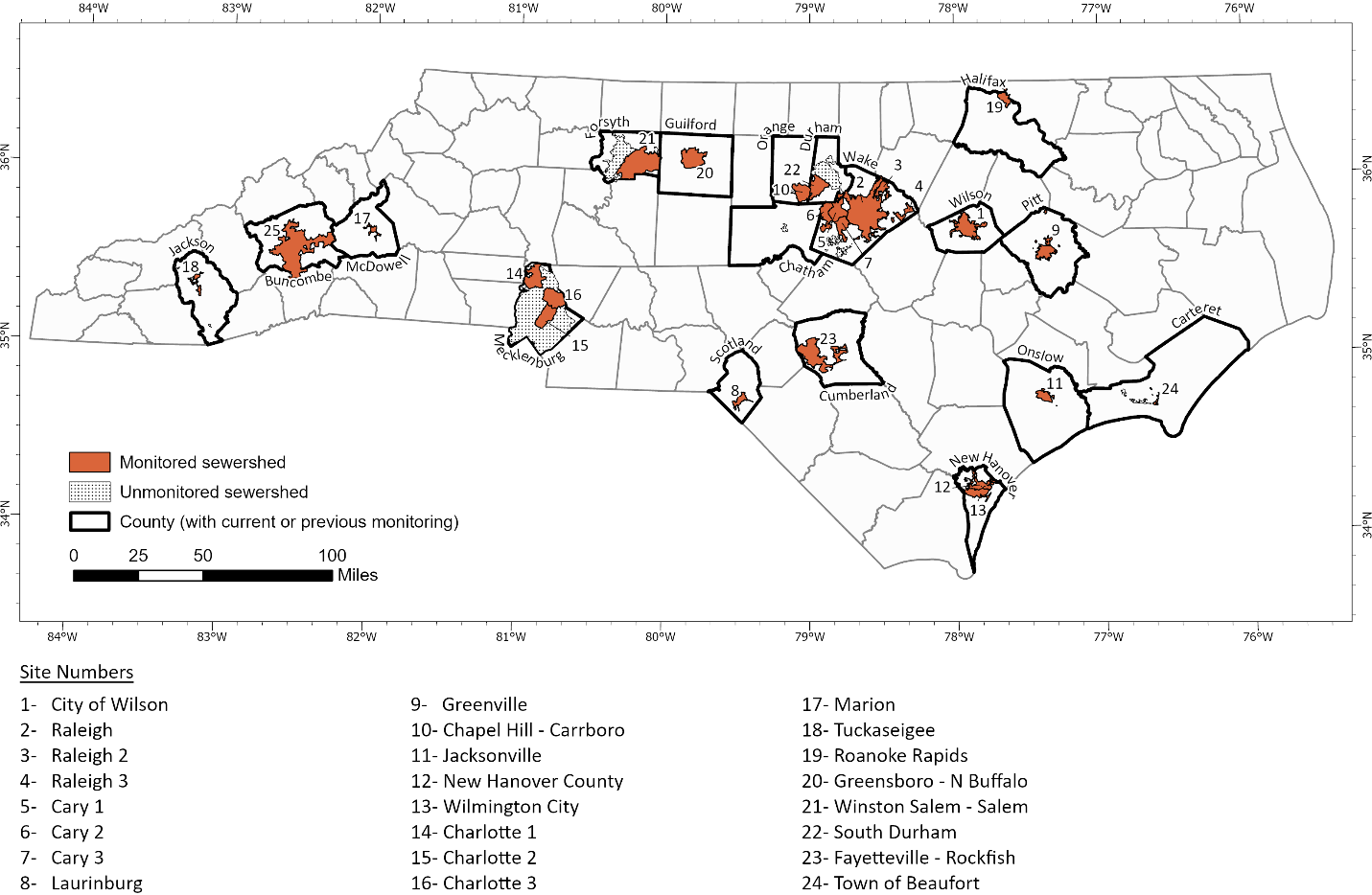

Note: Sewersheds reflect the service areas of wastewater treatment plants. Monitored sewersheds are those that were participating with the North Carolina Wastewater Monitoring Network as of June 2022. More than one sewershed was monitored in Mecklenburg County (n=3), New Hanover County (n=2) and Wake County (n=6).
